# Infection Units: A novel approach to the modeling of COVID-19 spread

**DOI:** 10.1101/2021.05.01.21256433

**Authors:** Jose C. Merchuk, Francisco García-Camacho, Lorenzo López-Rosales

## Abstract

A novel mechanistic model describing the rate of COVID-19 spread is presented, that differs conceptually from previously published deterministic models. One of its main characteristics is that the pool of infected people is not assumed to be homogeneously mixed, but rather as a passage into which individuals enter upon contagion, move within it in a plug-flow manner and leave at recovery, within a fixed time period. So, the present model differs conceptually in the way it describes the dynamics of infection. An ‘infection unit’ is defined as the amount of COVID-19 virus that generates contagion, if it reaches a susceptible individual. This model separately considers various pools: symptomatic and asymptomatic infected patients; three different pools of recovered individuals; pools of assisted, hospitalized patients; the quarantined and, finally, those who died from COVID-19. The transmission of the disease from an infected person to others is described by an ***infection rate function***, while an ***encounter frequency function*** modulates the frequency of effective encounters between the infected and the susceptible. The influence of the model’s parameters on the predicted results is presented. The effect of social restrictions and of quarantine policy on pandemic spread is shown. For model calibration, a set of experimental data is used. The model enables the calculation of the actual behaviour of the studied pools during pandemic spread.

## 1. Introduction

It seems that COVID-19 is one of the hardest health problems that humanity has had to deal with throughout its history, not so much because of the severity of the disease, nor its rate of spreading, but because of its global impact as the most rapidly widespread pandemic. This is the case due to the 21^st^ century combination of accessible, advanced transportation technology and the large volume of international travel for both business and pleasure—a blatant feature of modern consumer societies. Currently, almost all the countries in the world are engaged in trial-and-error processes, in which sanitary measures (including vaccination rate) are competing with economic activities of all kinds in battles between public health management and sustainable population maintenance (Ceylan, 2020). There is need of a macro-model that describes, as closely as possible, the whole of this complex issue (Acemoglu et al., 2020; Alvarez et al., 2020). Within such a macro-model, the modelling of the epidemiologic aspect and its particularities is of extreme relevance.

The number of research papers published on COVID-19 is vast, and their scope is broad (Fraser et al., 2021). Sometimes, the spread of a pandemic has tendencies that seem random. Therefore, statistical methods have been applied to predict the spread of such diseases, since they take multiple factors into account by means of time-series models, multivariate linear regressions, grey forecasting models, backpropagation neural networks, etc. However, the aforementioned statistical tools seem to be insufficient for analyzing pandemic randomness, and these models are difficult to generalize, as noted by Ceylan (2020). He claims that the COVID-19 prevalence in several European countries may be described using variants of an autoregressive integrated moving average (“ARIMA”) model, a time-series-type model. Wu et al. (Wu et al., 2020) simulated the expansion of COVID-9 across the most populated Chinese cities connected by airlines, using what they called ‘a metapopulation model’ with the SEIR variables (***S***usceptible, ***E***xposed, ***I***nfectious, ***R***ecovered). The basic calculations applied Markov chain Monte Carlo methods. Models of this type are called ‘agent-based’ models because they focus on the movement of/and contact between individuals. They require laborious calculations that provide a geographical aspect to pandemic spread. Hunter et al. made a detailed comparison of equation-based models versus agent-based models (Hunter et al., 2018). Recently Tsori and Granek (Tsori & Granek, 2020), among others, commented that most of the deterministic models are of the SEIR-type; they also stressed the fact that a ‘perfect mixing’, that is ‘total homogeneity’ in each of the pools, is always assumed. They pursue mitigation of this limitation by formulating “a continuous spatial model based on nearest-neighbor infection kinetics,” which leads to a reaction-diffusion-type description of pandemic spread. This enables the description of the spatial spread of a pandemic; their results show the two-dimensional spread across an actual geographical map. Nevertheless, their model maintains the same mathematical format as the SEIR-type models by describing the infected (***I*)** pool as a mixed compartment, in the sense that an individual classified as ‘infected’ may leave this compartment independently of his/her ‘external age’, i.e., the time spent in it (Danckwerts, 1953); here, ‘age’ is defined, therefore, as the average time an infected individual spends in the ***I*** pool. (The significance of this point is explained below.) The Manenti et al. model describes the entire population of the world as a perfectly mixed batch reactor (Manenti et al., 2020) and reaches formulations that are equivalent to the classic SEIR or SIR models, as may be expected. Cao et al. proposed an improvement in the 6-compartment SEIR-type model that adds a pool of quarantined patients (Cao et al., 2020). They used a time-series analysis exponential smoothing method and the ARIMAX model, often used in statistical modeling to analyze changes that occur over time. A considerable improvement in the SEIR-type models was done by Ivorra et al. (Ivorra et al., 2020). They called it the *θ*-SEIHRD model, and it was based on their previously published Bi-CoDis model (Ivorra et al., 2015). Here, they added seroprevalence as a measured variable, which is a very important addition to a system with a scant number of measured variables.

In this work, we focus on the mechanisms of pandemic spread and present a deterministic model with a novel approach. Our model differs conceptually in the manner of description of the transmission dynamics of the disease, the key being the ‘external age distribution’ of the individuals exiting the ***I*** pool (Danckwerts, 1953).

### 1.1 The flow dynamics of the infected (I) population pool

Here, we try to focus on one basic concept implied in the SEIR formulation and propose an alternative. The SEIR-inspired models, in all their variants, collide with the basic observation that the duration of this illness is an almost constant number of days (Manenti et al., 2020).

The well-known SEIR models describe distinct pools of the susceptible (***S***), exposed (***E***), infectious (***I***), and recovered (***R***), and sometimes additional pools--all of them defined as completely mixed. In a completely mixed continuous system, the systemic response to a pulse disturbance will always have a bell shape (in the case of an ideal instantaneous pulse, to a descending exponential), as described in basic textbooks (Levenspiel, 1972). This bell shape represents the ‘age distribution’ within the compartment; consequently, the ‘age’ of the individuals leaving the ***I*** pool will show a wide distribution. There would be individuals who stay in that pool a very short time, near zero, and others who stay in the pool a very long time, near infinite. This blatantly contradicts our knowledge about the behaviour of COVID-19 and other Coronaviruses that produce sicknesses with quite defined durations. Therefore, SEIR-type models fail to properly predict this viral infection mechanism, especially for shorter time periods. The ***I*** pool, as it will be defined here, has an inlet of individuals from the pool of the susceptible, ***S***, and outlets to other pools, but has a completely different behaviour. In the terminology of process engineering, the exhibited behaviour is called ‘a plug flow system’, resembling a conveyor belt, transporting infected individuals. The main characteristic defining such a system is its population dynamics, i.e., the homogeneity of the ‘age’ of the individuals leaving the compartment. The ‘age’ of the elements leaving the system cluster around a certain mean ‘residence time’, ***t***_***r***_, with relatively small variance. In practice, it is well known that a COVID-19-infected individual stays as such for a finite and quite defined period of time only, as estimated in the literature on the basis of experimental findings (Bar-On et al., 2020). Though there may be some individual variations in the case of COVID-19, this period of time seems to be consistently around 2 weeks (Liu et al., 2020). The state of the patient changes throughout this period and may lead either to recovery or to a more severe state and then, either to a full recovery or to death. The actual events during a normal ***I*** period take place along one clear timeline in an orderly manner. This is a basic characteristic of the illness, and the SEIR models fail to describe it. Such models may fit the dynamics of infection in the case of a population pool over long periods of time but cannot describe the short-term dynamics. Here, we present an alternative approach that overcomes this weakness.

## 2. An Alternative Approach

### 2.1. The I pool

A ‘plug flow model’ is a diametrically opposed alternative to the totally mixed compartments that characterize the SEIR-type models. In a plug flow system, all the elements that enter a compartment will leave it after residing in it a finite time, ***t***_***r***_, which corresponds to the term ‘serial interval’, commonly used in epidemiology (Last, 2001). In terms of our model, all the infected individuals will remain in this condition approximately ***t***_***r***_ days.

In an ideal plug flow situation, any change in the input at *t*=0 will produce the same (or similar) signal at the outlet, at ***t=t***_***r***_. In other words, the ‘age’ of each infected individual will increase steadily from 0 to ***t***_***r***_, which represents the period during which he/she was a member of the ***I*** pool. In practice, there will obviously be a certain distribution around the mean value but, in this version of the model, we disregard it.

For convenience in formulating this model, a dimensionless ‘residence time’ (***τ*=*t*/*t***_***r***_) in the ***I*** pool has been defined, as follows. The value of ***τ*** varies between 0 (input) and 1 (output), running parallel to time ***t***. It is assumed that all the infected individuals contained within the ***I*** pool at any time (***t*>*t***_***r***_) behave similarly after being infected. The period of ***t*<*t***_***r***_ must be treated somewhat differently, as will be shown below.

Based on the definition of ***τ***, Fig. 1 schematizes how individuals, with different ***I*** ages, are located throughout the entire ***I*** pool (top of the figure) **at a given time *t***. At the bottom of Fig. 1, a single individual is shown in transit through the course of the illness (**0<*τ*<1**). For any ***t*>*t***_***r***_, a record of all the data in the entire ***I*** pool would show the whole length of the passage or conveyor belt that represents the pool of the infected, occupied by infected individuals of increasing ‘age’ (‘residence time’ in the ***I*** pool). In other words, at the pool inlet (***τ*=0**), ***I***(***t***) newly infected would be entering, while the infected individuals located within **0<*τ*<1** would be those who entered the pool at an earlier time (***t*-*τ*·*t***_***r***_). The ‘oldest’ individuals, after ***t***_***r***_ days of illness, are found at the exit, ***τ***=1, leaving the ***I*** pool. For the special case, in which the time elapsed from the start of the pandemic is exactly ***t*=*t***_***r***_, those individuals who are just about to leave the ***I*** pool (at ***τ*=1**) will be those first, susceptible individuals who had just become infected and started their infection period at ***t*=*τ*=0**.

**Figure 1.**
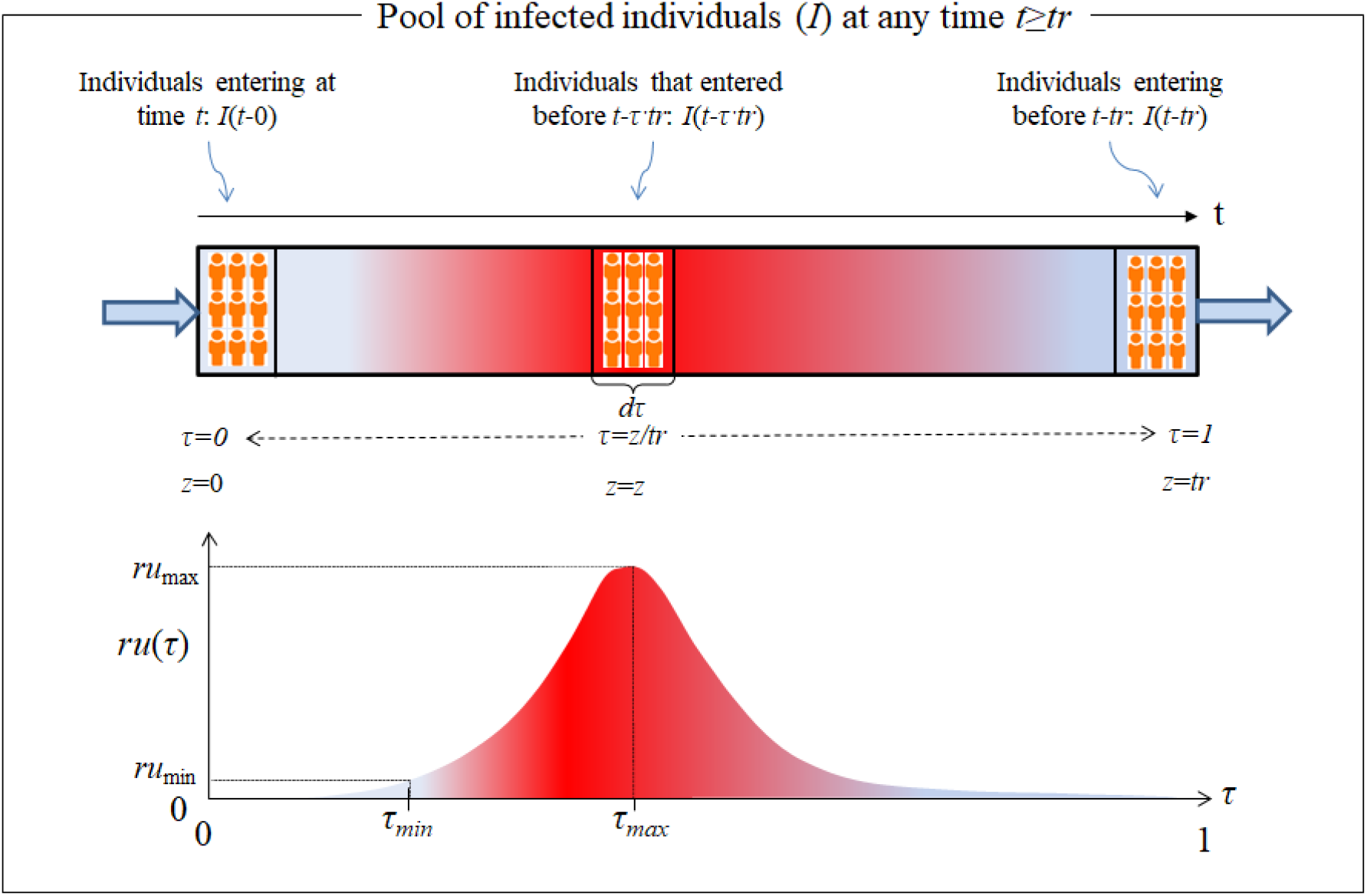
Top: The ***I*** pool as it behaves over time, ***t***. Bottom: The transit of a single patient through the ***I*** pool is delimited from the time of infection (inlet at ***τ*=0**) to the end of the sickness (outlet at ***τ*=1**).

There is another particular case that occurs when the time elapsed from the beginning of the pandemic is ***t*<*t***_***r***_. In this case, ‘patient zero’ is located on the ‘moving sidewalk’ at ***τ*=*t*/*t***_***r***_ and has not yet left the ***I*** pool, the end of which is located at ***τ*=1**. As result, the ‘moving sidewalk’ is empty in the interval [***τ***,**1**]. Other than this special case, all patients start the transit through the ***I*** pool at ***τ*=0** and exit at ***τ*=1**.

Before describing the ***I*** pool, we must introduce our definition of the ‘***infection unit***’ **(*U***) – the amount of COVID-19 virus released by an infectious individual in any way or form having the capacity to cause the conversion of susceptible (***S***) individuals into infected (***I***) ones. This definition is inspired by the ‘photosynthetic unit’ concept, **PSU**, that is widely accepted in the area of photosynthesis modelling (Megard et al., 1984; Merchuk et al., 2019; Prézelin, 1981). Initially, the PSU concept seems intangible, yet it produces excellent practical results. We consider that the same may be true for our viral ‘infection unit’ concept, ***U***. An infected individual moves through the ***I*** pool, producing ***U*** at a certain rate, ***r***_***u***_***(τ)***. This rate is only the result of the disease incubating in his body during his/hers transit through the illness (from ***τ***=0 to ***τ***=1) and will, therefore, not be affected by any of the variables in the model. A shifted Gaussian function, as shown in the lower part of Fig. 1, was chosen arbitrarily to represent ***r***_***u***_***(τ)***. Other functions may be used without changing the underlying concept.

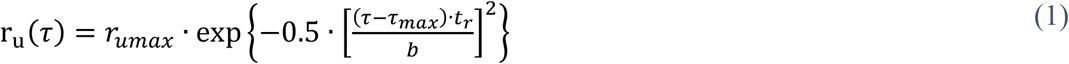

The curve plotted in Fig. 1 concurs with the actual behaviour of the virus. Once the invading virus enters the body, it requires a certain amount of time to locate and penetrate its target cell, before activating its cellular machinery to produce the first generation of native virus. The durations of some of those steps have been reported for other viruses (Zhang et al., 2020). Then, additional time (an ‘eclipse phase’) necessarily follows, during which more cells must be infected and finally reach ***τ***_***min***_, the point in time at which the actual dissemination of the virus from the ill person’s body takes place. At this stage, the infection may finally be diagnosed. Since ***r***_***u***_***(τ)*** starts at zero and at the end of the infectious period it will once again be zero or close to it, it is obvious that, at some point (***τ*=*τ***_***max***_), there will be a peak in viral dissemination rate (***r***_***umax***_), followed by a ***r***_***u***_***(τ)*** decrease. Here, it is assumed that there is a certain minimum rate of ***U*** production, below which there is no infectivity (***r***_***umin***_), as shown in Fig. 1. At the end of the period (***τ*=1**), the patient recuperates and passes into the *R* pool, unless his/her health deteriorates. This last situation will also be considered below.

The generation of ***U*** is necessary, but on its own, it is not enough to transmit an infection. For transmission to occur, the presence of a susceptible individual is also required. In the SEIR models, the presence of susceptible individuals is permanent, as implied by the assumed kinetic form of the infection rate, where the product ***I*** times ***S*** appears. As long as ***S*** is not null, some degree of infection occurs. Nonetheless, in practical, interpersonal encounters, an infected individual will not always be within physical infectivity range, nor will the quantity of the ***U*** released by an infectious person be enough to effectively transmit this viral disease. This fact is expressed by means of our ***encounter frequency function, f*(*t*)**, where ***t*** is the timeline along which the pandemic progresses. We will focus on the case of a typical, <u>undetected</u>, asymptomatic person with COVID-19.

The transmission of this infection via individuals who carry the virus, but are asymptomatic, is a characteristic of COVID-19 and was seldom seen in the SARS and MERS outbreaks. As long as the disease has not yet been diagnosed, no special insolation measures are taken. It may be assumed that there are different types of encounters between infected and susceptible members in a community related to lifestyles, usual daily routines, essentially repetitious in nature (e.g., work, study, shopping, hobbies, regular schedules and means of transportation). The importance of such regular routines has been recognized Backer et al. (Backer et al., 2020). There are also certain non-routine activities, often involving a larger number of people (e.g., weddings, parties, dinners, etc.). The abovementioned “super-spreading events” can easily be included here in addition to individual singularities but here, for the sake of simplicity, we intentionally present only the repetitive daily encounters as periodic functions. One of the simplest formulations for this ***encounter frequency function, f*(*t*)**, is:

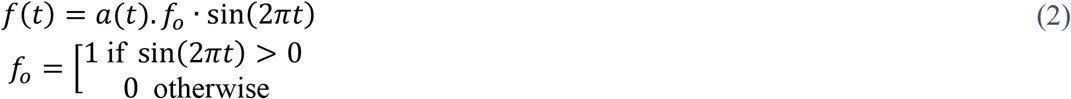

In expression (2), the influence of the restrictions imposed on social contacts (‘social distancing’) and their impact on the behaviour of the population may be accounted for in ***a*(*t*)**, which is instrumental in the modulation of the infection rate constant (***K***_***SI***_). The need for this modulation has already been indicated in the literature (Canabarro et al., 2020; Loli Piccolomini & Zama, 2020). We adopted the following function based on another recent report (Prasad, 2020):

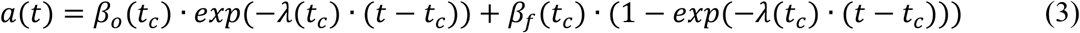

where it is assumed that, once a restriction has been imposed by the authorities and it starts on a certain fixed day (***t***_***c***_), the level of social contact will decay at a rate of ***λ*** from a starting value ***β***_***o***_, tending towards a limit value of ***β***_***f***_. Obviously, the greater the severity of the restrictions, the higher the ***λ*** value and the lower the ***β***_***f***_ value. The Eq. (3) above may be used for ***i*** consecutive constraints; with ***λ***_***i***_ values for each interval ***t***_***ci***_**-*t***_***ci+1***_, on which the ***i*** restriction is set. As previously detailed (Loli Piccolomini & Zama, 2020), to improve the flexibility of Eq. (3), the integration interval may be split into sub-intervals with widths that do not have to coincide exactly with the days on which the restrictions were decreed.

It should also be taken into account that not every infected person in the ***I*** pool will transmit the disease. In this paper, we assume that all <u>diagnosed</u> patients (***I***_***d***_), are under proper care, completely isolated, in which case ***f*(*t*)=0** and they cease to be contagion factors; the same applies to all <u>assisted</u>, hospitalized patients. However, infected individuals who have not yet been identified as such because they are not showing blatant symptoms, are considered in the present version of this model as being the only factors of pandemic transmission.

The ***U*** defined above are generated at the rate of ***r***_***u***_***(τ*)**, produced during the evolution of the disease within the body of each patient during his/her passage through the illness (from ***τ*=0** to ***τ*=1**); as such, this rate is not affected by any of the variables in the model. By definition, ***ru*** represents the rate of generation of the ***U*** by a single infected individual. In order to calculate total ***U*** generation, the number of infected individuals is required. The number of <u>undetected</u> infected persons at time ***t*** who are at stage ***τ*** of the illness is ***I***_***nd***_**(*t*-*τ*·*t***_***r***_). It is convenient to define the total number of ***U*** generated by individuals in the state of infection **τ** at a given time ***t*** as ***U***(***t***,***τ***):

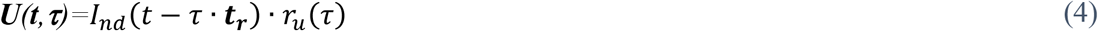

Here, ***I***_***nd***_**(*t-τ·t***_***r***_) is the number of ***I***_***nd***_ who entered the ***I***_***nd***_ pool at an earlier time (***t*-*τ*·*t***_***r***_) and, as a result, they have an ‘infectivity rate’ of ***r***_***u***_**(*τ*)** at a given time ***t***, where **τ** is the dimensionless ‘residence time’ (***t***_***r***_) in the ***I***_***nd***_ pool, that runs along time, taking values from **τ=0** to **τ=1**, as previously explained. Note that Eq. (4) renders the ***U*** production rate of infected individuals at time ***t*** in state **τ**. Multiplying by the ***encounter function***, we get the number of ***U*** able to transmit infection. Integrating over the range of **τ**, we obtain the total rate of infection at any time, ***t>t***_***r***_, within the *I*_*nd*_ pool:

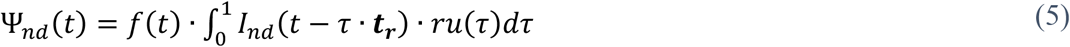

Equation (5) shows that, for the calculation at a time ***t***, we must refer back to past values of ***I***_***nd***_**(*t*)** (before ***t***), following Eq. (4). (The precise numerical calculation procedure is detailed later on in this paper.) Note that, for ***t*<*t***_***r***_, the number of infected individuals coincides with the initial condition ***I***_***nd***_**(0)**. For the interval **0<*t*≤*t***_***r***_, we have modified Eq. (5) as follows:

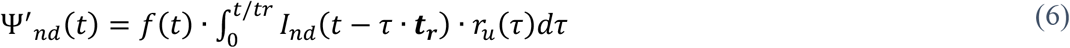

### 2.2 Description of the model

The history of a single member of the entire ***I*** pool, from the time of his/her infection (***τ=0***), is presented here. At this stage, it is important to note the distinction between <u>undetected</u> individuals (asymptomatic), who are unaware of their medical conditions and to whom no social-distancing restrictions are yet applied, and those <u>diagnosed</u> patients, identified as having COVID-19, who are assumed to be under quarantine. The general scheme of the model may be seen in Fig. 2 below.

**Figure 2.**
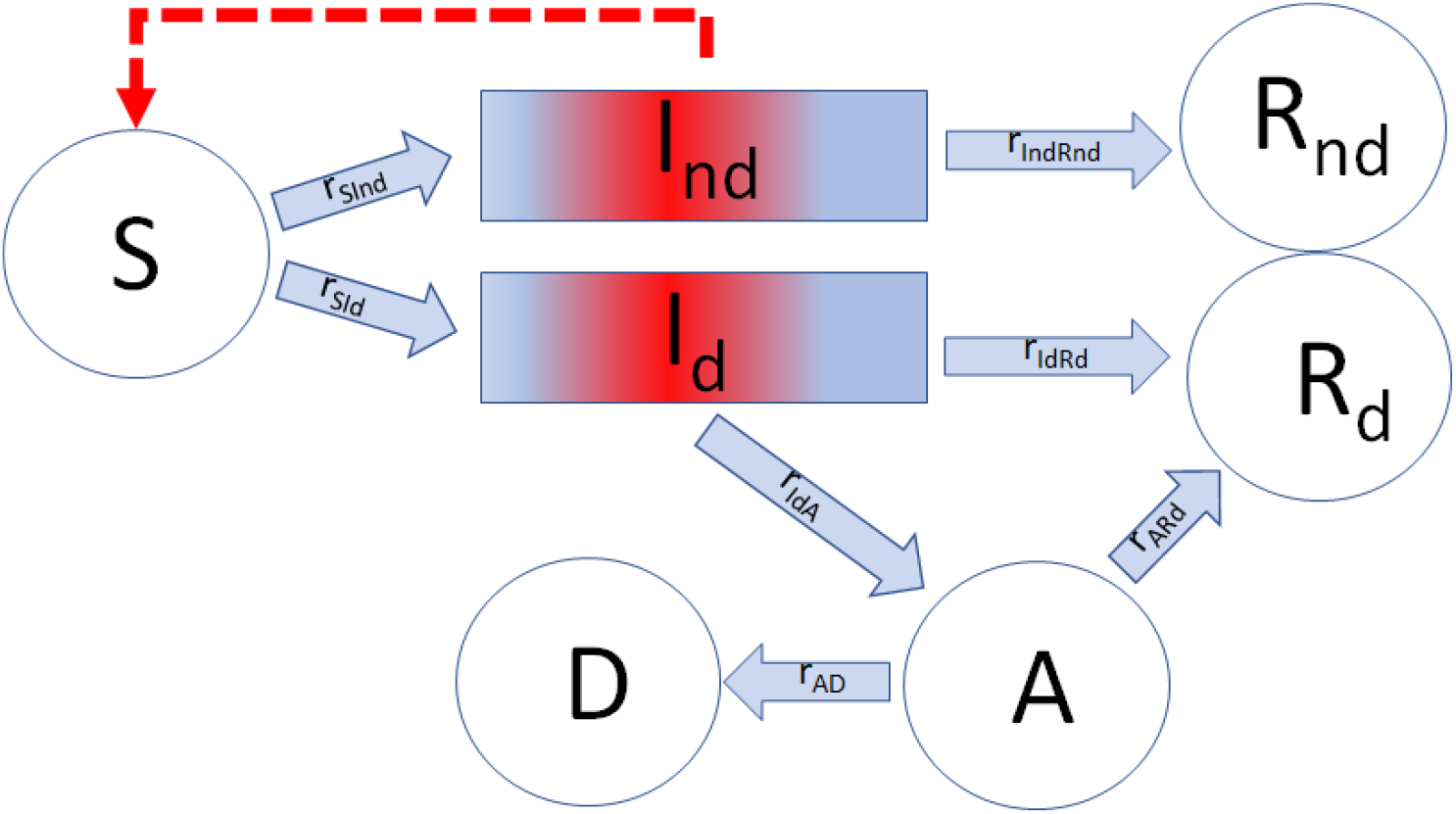
***I***_***d***_ is the pool of <u>diagnosed</u> patients; ***I***_***nd***_ is the pool of <u>undetected</u> individuals; ***R***_***nd***_ includes those <u>undetected and recuperated</u>; ***R***_***d***_ includes those <u>diagnosed and recuperated</u>; ***A*** - the <u>assisted</u>, hospitalized patients; ***D*** - the <u>dead</u>; ***S*** - those <u>susceptible</u>. The broken lines indicate the action of the ***U*** that catalyze the conversion of ***S*** into ***I***. The ***r***_***i***_ represents the flux rate between the different pools.

In Fig. 2, the rectangles indicate that the individuals in two ***I*** pools behave as described in Fig. 1 in regard to time dependence, ***t***_***r***_, while the circles symbolize other pools, in which the exit time, ***t***, is not dependent on the ‘residence age’; that is, the sub-population is ‘perfectly mixed’.

That fraction of infected individuals who are not diagnosed, either because they were asymptomatic or because the symptoms were mild and went unnoticed, constitute the pool, ***I***_***nd***_. They are, however, <u>contagious</u> and can infect the susceptible population (***S***) in accordance with the ***U*** dissemination mechanism described above. All the undiagnosed individuals in the ***I***_***nd***_ pool recover after a time of ***t***_***r***_, when they become members of the corresponding pool of the diagnosed and recovered patients, ***R***_***nd***_. The remainder of the total infected, ***I***_***d***,_ develop characteristic symptoms after the period of incubation and are then isolated; thus, we assume that they no longer serve as factors in contagion. After ***t***_***r***_, when the ***I***_***d***_ recuperate, they enter the pool ***R***_***d***_. An alternative case is when a patient suffers a complication during his/her illness (related or not to pre-existing problems) and the patient’s health deteriorates further, requiring some kind of intensive treatment—thus passing to the assisted, hospitalized (***A***) pool. The point in the patient’s history, at which this change in status may possibly take place, is symbolized as ***τ***_***IdA***_. The patients in the ***A*** pool either recuperate and transit into ***R***_***d***_ or, contrarily, their condition deteriorates to a state of death, when they join the ***D*** pool, having died of COVID-19.

Our model describes the mechanism of COVID-19 dissemination as a series of mathematical expressions, symbolized by the different arrows in Fig. 2.

### 2.3. Balance in the *I*_*nd*_ *pool*

We define the total number of infected, ***I***, as the sum of all the diagnosed and undetected infected individuals:

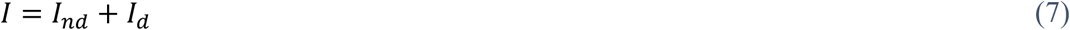

Step *S*→*I*, the contagion is itself, is responsible for the generation of all the infected, ***I***_***nd***_ and ***I***_***d***_, at rate ***r***_***SI***_. This step is catalyzed by the ***U*** generated within the ***I***_***nd***_ pool, in accordance with the mechanism described above. Therefore, ***r***_***SI***_ can be presented as Eq. (5):

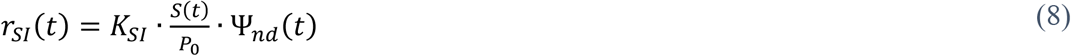

The term 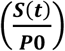 is the fraction of susceptible individuals in the population and gives physical meaning to the ***encounter function, f*(*t*)**. The constant ***K***_***SI***_ characterizes the infectivity of the virus, though it may be somewhat different for different genetic variants of the same virus. The overall change in the number of individuals in the ***I***_***nd***_ pool represents the balance of the fluxes, ***S*→*I*** (influx) and ***I***_***nd***_**→*R***_***nd***_ (outflow). A fraction of the ***I*** generated corresponds to the ***I***_***nd***_, the rest to the ***I***_***d***_.

Defining the ratio ***ϕ*** as:

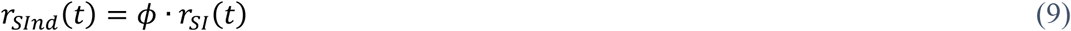

it is obvious that the rate of the undetected, infected (***I***_***nd***_), moving to the pool of the undetected, recuperated, ***r***_***IndRnd***_, will be similar to the inpu, ***r***_***SInd***_ but shifted by ***t***_***r***_ days, thus:

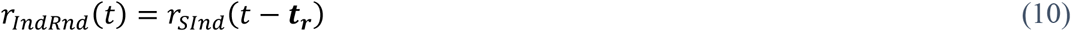

This means that the first individuals to leave the ***I***_***nd***_ pool and reach the ***R***_***nd***_ pool are those who first entered the ***I***_***nd***_ pool ***t***_***r***_ days earlier. This assures the simple fact that no patient will recuperate before a time ***t***_***r***_ from the beginning of the process (***t*=0**), as usually observed in practice. A similar delay will take place in the transit from ***I***_***d***_ to ***R***_***d***_. This delay is not described by the SIR-type models.

In our model, the rate of change of ***I***_***nd***_ is given simply by:

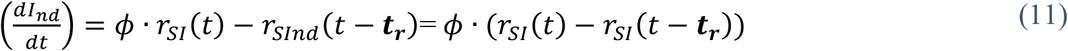

Equation (11) simply states that the accumulation of the infected, ***I***_***nd***_, is equal to the difference between those generated and those exiting that pool to the pool of the recuperated, ***R***_***nd***_.

### 2.4 Balance within the *I*_*d*_ *pool*

The population in the ***I***_***d***_ pool is kept balanced by the influx of ***S*→*I***_***d***_ (***r***_***SId***_) and two outflows, ***I***_***d***_**→*R***_***d***_ (***r***_***IdRd***_) and ***I***_***d***_**→*A*** (***r***_***IdA***_). However, a considerable simplification may be obtained by accepting the abovementioned assumption (derived from Eq. (9)) about the relationship presented below (in Eq. (12)), that takes advantage of seroprevalence information:

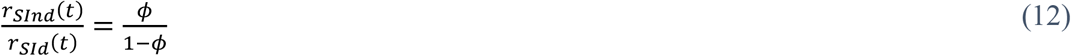

letting us write:

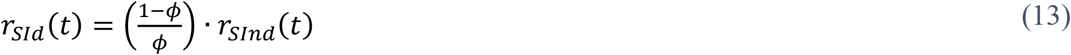

It is intuitive that an infected patient will not become gravely ill immediately after infection, since a certain period is required before the patient may reach a state of severe illness. The rate of transit (***r***_***IdA***_) from ***I***_***d***_ to ***A***, the pool of assisted, hospitalized patients, applies to some of the infected individuals, ***I***_***d***_ (***t*-*τ***_***IdA***_**· *t***_***r***_), at an ‘age’ ***τ***_***IdA***_ in the ***I***_***d***_ pool. The range of possible complications in this disease is extremely wide, since it depends on the patients’ previous health conditions. It would be extremely difficult to contemplate all the possibilities and, therefore, we assume the simplest mathematical form, and ***r***_***IdA***_ is written as:

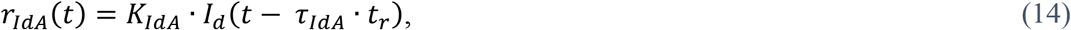

where it has also been assumed that the rate of patients exiting to pool ***A*** is proportional to ***I***_***d***_**(*t*-τ**_***IdA***_ **· *t***_***r***_). It seems that this is the likely point at which this disease may lead to complications in a fraction of the patients at the ‘age’ of **τ**_***IdA***_. This roughly agrees with reported field observations (Novack, 2020). In the special case of ***t*<τ**_***IdA***_***·t***_***r***_, no patient will have yet exited to the assisted, hospitalized *A* pool; such an event can only occur after ***t*=τ**_***IdA***_***·t***_***r***_. The output from the ***I***_***d***_ pool to the ***R***_***d***_ pool, consisting of the diagnosed, recuperated patients, is written as:

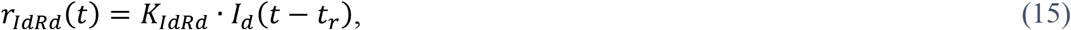

where it has been assumed that the exit rate to pool ***R***_***d***_ is proportional to the number of patients ***I***_***d***_**(*t-t***_***r***_), and that no patient can recover before ***t***_***r***_ days from the initial time of infection (***t*=0**). Therefore, the mathematical formulation of this balance is presented as:

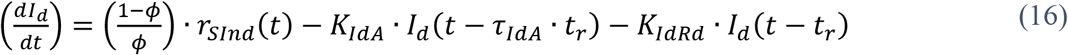

### 2.5 The rest of the pools

The formulations for the rest of the pools are much simpler. There are no fixed time-frames of which we are aware for severely ill patients who remain in pool ***A***. The pools ***R***_***d***_, ***R***_***nd***_, and ***D*** grow monotonically over time, while pool ***S*** can only shrink, since we do not consider births nor deaths as causes of COVID-19.

The balances for the recuperated patients’ pools, ***R***_***d***_ and ***R***_***nd***_ (both these pools are considered here to be resistant to COVID-19) involve the following fluxes: ***I***_***nd***_**→*R***_***nd***_ **(*r***_***IndRnd***_); ***I***_***d***_**→*R***_***d***_ **(*r***_***IdRd***_); and ***A*→*R***_***d***_ **(*r***_***ARd***_). Flux ***r***_***IndRnd***_ (from Eq. (10)) represents undetected, infected patients who have recuperated; flux ***r***_***IdRd***_ (from Eq. (15)) represents diagnosed, infected patients who have recuperated; and ***r***_***ARd***_ represents those who recuperated after a period of assisted, hospital treatments. Our mathematical expression for ***r***_***ARd***_ follows a simple first-order law:

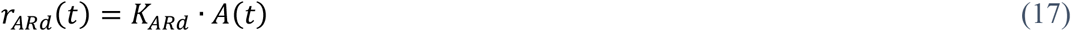

The balances for pools ***R***_***d***_, ***R***_***nd***_ and ***R*** are formulated as:

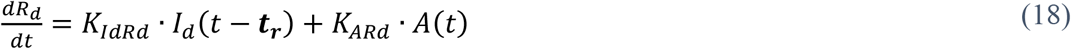

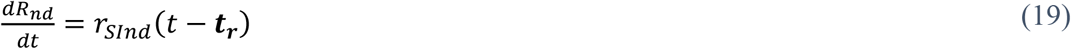

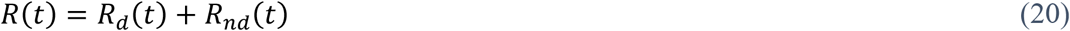

Similarly, the balance of the assisted, hospitalized patients’ pool, ***A***, is written as:

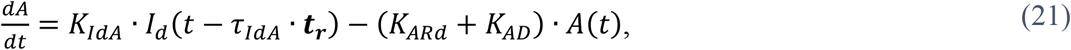

where the first term on the right-hand side of Eq. (21) represents the flux into pool ***A*** coming from pool ***I***_***d***_ and the second term is the sum of the recuperation and death rates of the assisted, hospitalized patients.

The rate of change in the balance of COVID-19 mortality assumes that all the patients who reach pool ***D*** previously passed through pool ***A***. It is written as:

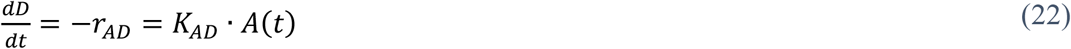

The balance of the susceptible patients’ pool ***S*** is written as:

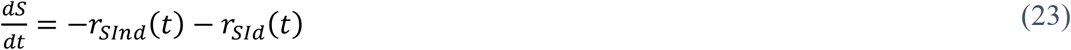

In Eq. (23), the first and the second terms represent the outflows from the pool ***S*** to two types of infected pools, ***I***_***nd***_ and *I*_***d***_, respectively. Recall that neither births nor deaths, other than those due to COVID-19, are considered in the present model. Thus, the total population, *P*_*0*_, remains constant:

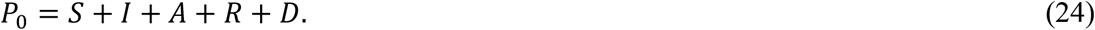

Since, in our model, natality and mortality other than from COVID-19 were not taken into account, the use of Eq. (24) makes redundant one of the balances presented in this model.

## 3. Model calibration

The hitherto described model (see scheme in Fig. 2) consists of a set of delay differential (DDE) and algebraic equations numbered from (1) to (24). In order to perform a model calibration and simulations thereof, we employed the MATLAB solver DDE23 with constant delays to integrate the system of DDEs. Our system is positive, since all the state variables have non-negative values for **t≥0**. The constant delays considered in the current formulation of the model were essentially two: ‘residence time’ (***t***_***r***_) in the ***I*** pool and ‘residence time’ (***t***_***r*·**_***τ***_***IdA***_) of those individuals shifted into pool ***A***. Other constant delays are those corresponding to the number of subintervals into which the limits of the integrations in Eqs. (5) and (6) are divided.

In order to estimate the parameters, data from the early evolution of COVID-19 in Italy (Giordano et al., 2020) was used, as an example of the application of the “Infection Units Model.” Table 1 shows the set of fitting parameters and the initial conditions for the DDEs used. We fit the solution of the set of DDEs to the measured data from the cumulative diagnosed, infected (**Σ*I***_***d***_), recovered ***R***_***d***_ and deceased ***D*** populations, relative to 35 days, from February 21, 2020 to March 26, 2020 (Giordano et al., 2020). The parameters of our model were calibrated by minimizing the following multi-output function, based on the average relative root mean squared error (aRRMSE), as elsewhere described (Borchani et al., 2015):

**Table 1.**
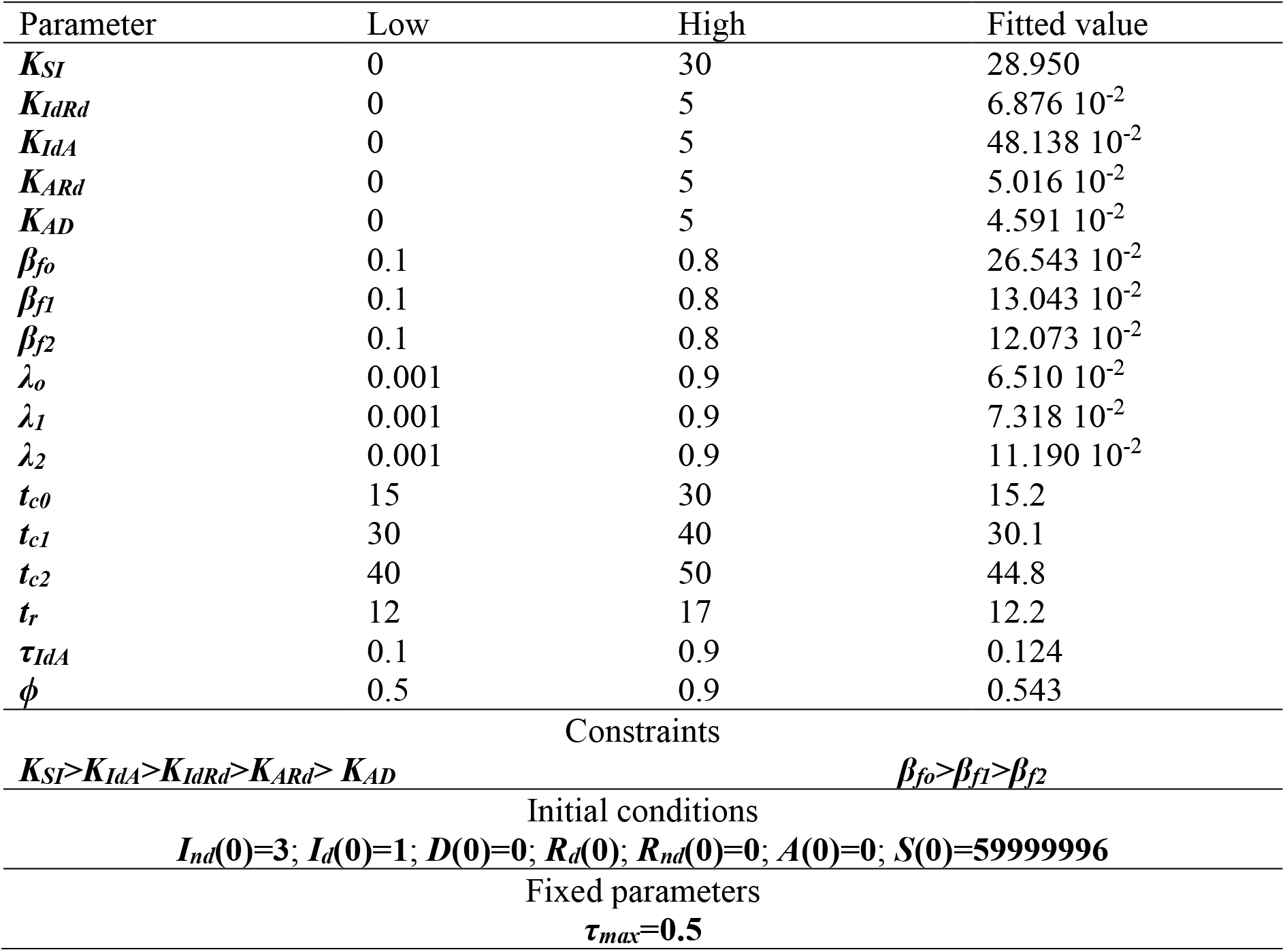
Displays a summary of the parameters identified with the Genetic Algorithm for the early development of the COVID-19 in Italy. The range of the boundary values (low and high), initial conditions, constraints, and fixed parameters during the optimization process are listed.

| Parameter | Low | High | Fitted value |
| --- | --- | --- | --- |
| $K_{SI}$ | 0 | 30 | 28.950 |
| $K_{IdRd}$ | 0 | 5 | $6.876 \cdot 10^{-2}$ |
| $K_{IdA}$ | 0 | 5 | $48.138 \cdot 10^{-2}$ |
| $K_{ARd}$ | 0 | 5 | $5.016 \cdot 10^{-2}$ |
| $K_{AD}$ | 0 | 5 | $4.591 \cdot 10^{-2}$ |
| $\beta_{fo}$ | 0.1 | 0.8 | $26.543 \cdot 10^{-2}$ |
| $\beta_{f1}$ | 0.1 | 0.8 | $13.043 \cdot 10^{-2}$ |
| $\beta_{f2}$ | 0.1 | 0.8 | $12.073 \cdot 10^{-2}$ |
| $\lambda_o$ | 0.001 | 0.9 | $6.510 \cdot 10^{-2}$ |
| $\lambda_1$ | 0.001 | 0.9 | $7.318 \cdot 10^{-2}$ |
| $\lambda_2$ | 0.001 | 0.9 | $11.190 \cdot 10^{-2}$ |
| $t_{c0}$ | 15 | 30 | 15.2 |
| $t_{c1}$ | 30 | 40 | 30.1 |
| $t_{c2}$ | 40 | 50 | 44.8 |
| $t_r$ | 12 | 17 | 12.2 |
| $\tau_{IdA}$ | 0.1 | 0.9 | 0.124 |
| $\phi$ | 0.5 | 0.9 | 0.543 |
| Constraints |  |  |  |
| $K_{SI}>K_{IdA}>K_{IdRd}>K_{ARd}> K_{AD}$ | | $\beta_{fo}>\beta_{f1}>\beta_{f2}$ | |
| Initial conditions |  |  |  |
| $I_{nd}(0)=3; I_d(0)=1; D(0)=0; R_d(0); R_{nd}(0)=0; A(0)=0; S(0)=59999996$ | | | |
| Fixed parameters |  |  |  |
| $\tau_{max}=0.5$ | | | |

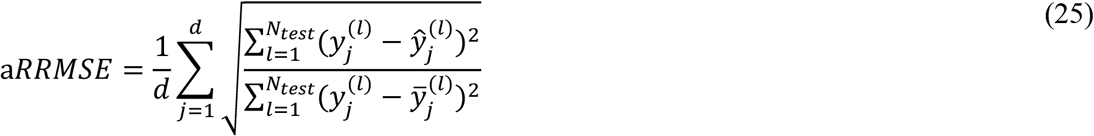

where ***y*** represents the experimental variable (i.e., **Σ*I***_***d***_, ***R***_***d***_ or ***D***); ***d*** is the number of experimental variables (i.e., **3**; **Σ*I***_***d***_, ***R***_***d***_ and ***D***); **N**_**test**_ is the size of the data set; 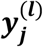 and and 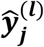 are the vectors of the actual and predicted outputs for the time vector, respectively, and 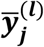 is the vector of average of the actual outputs. The aRRMSE automatically rescales the error contributions of each target variable (Borchani et al., 2015). Minimization of Eq. (25) was performed by using the MATLAB Genetic Algorithm Solver with the following options: (i) a crossover fraction of 0.8; (ii) “mutationadaptfeasible” as mutation function; (iii) “selectionroulette” as selection function; (iv) a population size of 500; and (v) 50 generations.

The available experimental data started from February 21, 2020, when the figures of the pandemic were low, but revealing: **Σ*I***_***d***_**=20, *R***_***d***_**=0** and ***D*=1**. If there had been a ‘patient zero’ (i.e., a single initiator, ***I***_***d***_**(0)=1**) of the COVID-19 outbreak, that individual must have emerged a few days earlier; as such, we assumed that this initial event happened around 15 days before. Thus, the integration interval was 50 days: a virtual first period of 15 days to capture the start of the pandemic outbreak, and a second period of 35 days that included the data from February 21, 2020.The initial conditions of this pandemic are displayed in Table 1 below.

Since, at that time, no seroprevalence data were available, ***ϕ*** is left here as a variable to be found by the optimization procedure. Neither were the data on ***A*** taken, due to the uncertainty regarding certain criteria, like the availability of free space (beds) and equipment for intensive treatment in the hospitals. The impact of the degree of compliance by the general population with the newly imposed regulations on ‘social distancing’ was taken into account. The Italian Government adopted new restrictive measures aimed at combating the spread of COVID-19. According to Italian announcements, the integration period (see Eq. 3 above) was divided into three phases, delimited by the corresponding ***t***_***c0-3***_ values in Table 1.

Consequently, three parameters were needed for ***λ*** and ***β***_***f***_ (see Table 1). The starting value, ***β***_***o***_, was fixed at 1.

The fitting of the model to the experimental data of Σ*I*_*d*_, *R*_*d*_, and *D* is displayed in Fig. 3; the fit is satisfactory (aRRMSE=0.0461). *A* was not taken as an input but as an output, to avoid the uncertainty in the reported data. This is related to the lack of homogeneity in medical criteria and the availability of hospital beds and medical devices in different locations. This fit renders the numerical values for the parameters shown in Table 1. Of special interest is the mean residence time of an ***I*** patient (serial interval), ***t***_***r***_**=12.2** days and the fraction of infected individuals who were undetected, ***ϕ* =0.543**. Both these values are within range of the reports available from multiple sources and strengthen the validity of our obtained results.

**Figure 3.**
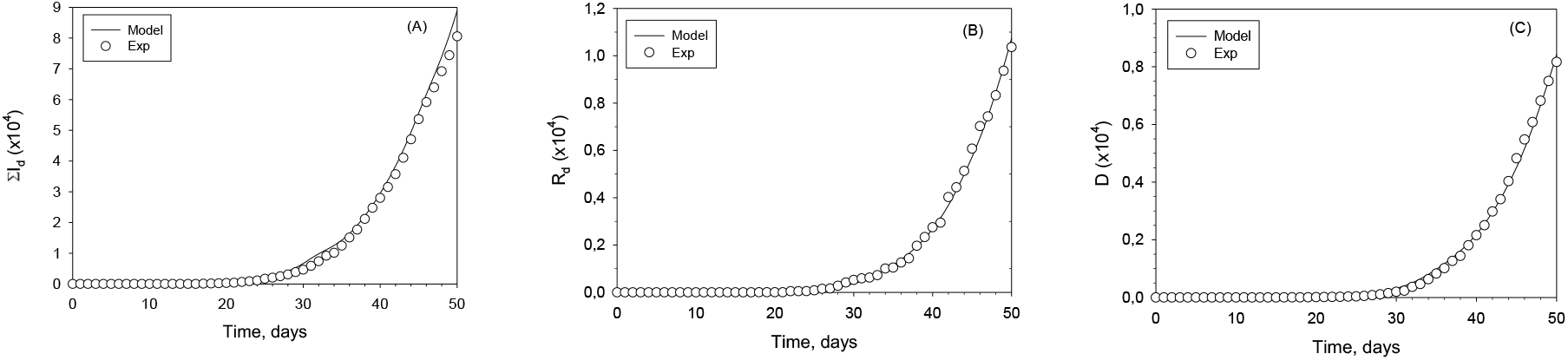
Infection Units Model fitted to Italy: (a) cumulative diagnosed, infected population, **Σ*I***_***d***_; (b) diagnosed, recovered population, ***R***_***d***_; (c) dead population, ***D***. Experimental data (cycles); modelled populations (continuous lines).

## 4. Model behaviour

In this section, we present numerical simulations that assess various characteristics of this model. To that end, simulations were run of our model extended to 200 days, with some modifications of the parameters from Table 1; each parameter was modified, one at a time, to reveal each one’s relative influence. These simulations were run with the parameters obtained with the fitted values listed in Table 1. The effect of the constant ***K***_***SI***_ on the predictions of the all variables (***S, I***_***d***_, ***I***_***nd***_, ***A, D, R***_***d***_ and ***R***_***nd***_) is presented in Fig. 4.

**Figure 4.**
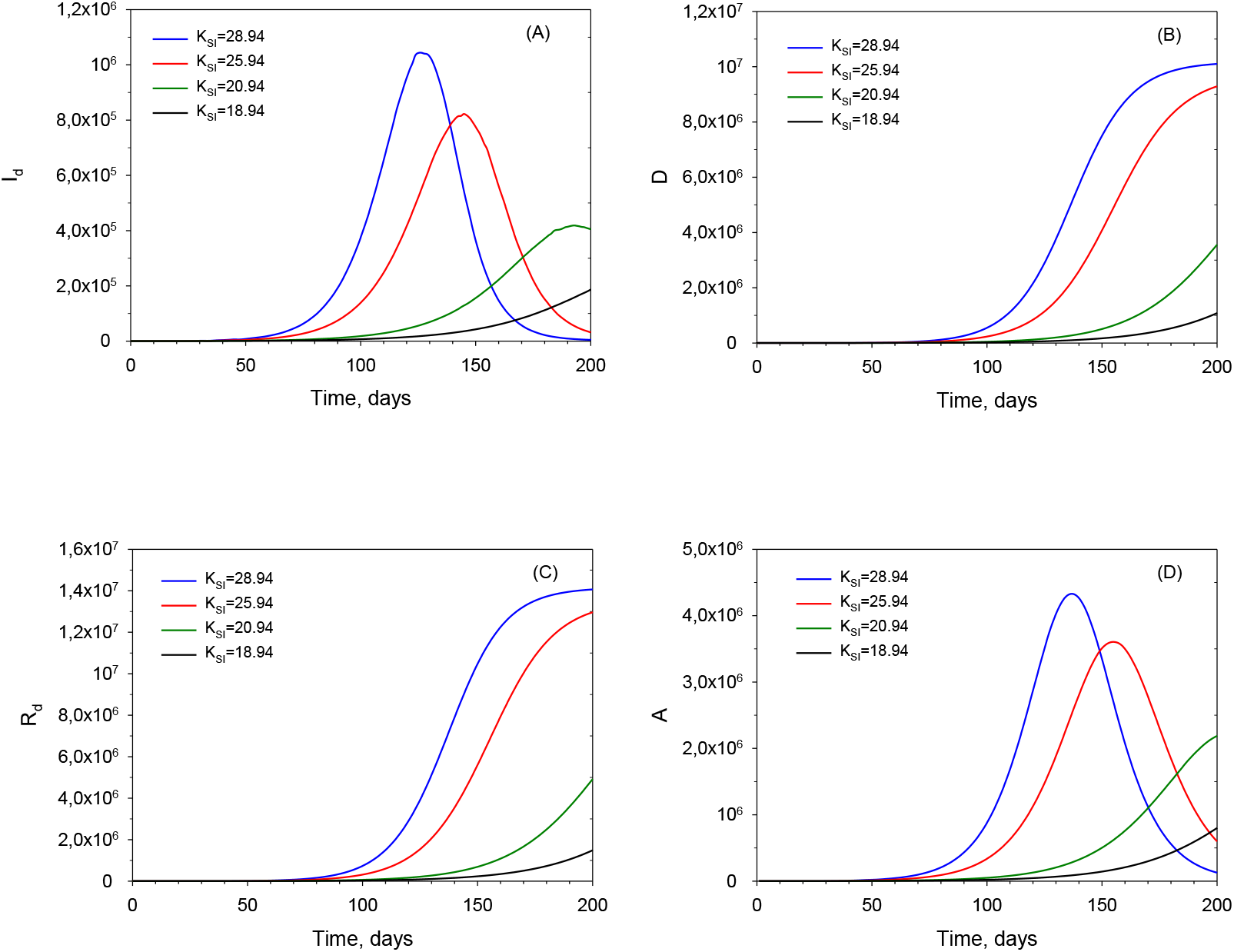
Calculated plots of those diagnosed, infected, dead and recuperated for increasing values of the basic infectivity coefficient, ***K***_***SI***_.

These plots include the time ranges and the field data used for the fittings, but should not be considered as extrapolations, rather as extensions of their ranges, for the purpose of qualitatively exploring the potential of our model. The constants in Table I are strictly valid for the time ranges on which they were based. For a more accurate simulation, research based on much more data over a longer time period should be carried out; such a study is currently beyond our means. The primary purpose of this study is to present our novel conceptual approach.

The constant, ***K***_***SI***_, which may be seen as an infectivity coefficient in the formulation of the rate of ***S*** decrease, ***r***_***SI***_, is one of the main factors affecting the performance of the model. Within the context of the COVID-19 pandemic, its variation range is rather limited to the altered infectivity of various the mutations. Nonetheless, a wider range of variation is shown in Fig. 4, in order to display the general characteristics and the potential of extension of our model to other viruses with a similar mechanism of dissemination. The profiles of the diagnosed, infected ***I***_***d***_, the dead ***D***, the assisted, hospitalized ***A*** and the diagnosed, recuperated ***R***_***d***_ are shown. Bear in mind that ***I***_***d***_***(t)*** is the number of instantaneous infected, and <u>not</u> the cumulative number that is published sometimes. The latter can be readily calculated from ***I***_***d***_***(t)***. As expected, the lowest values of ***K***_***SI***_ led to the failure of the virus to spread in the population. As ***K***_***SI***_ increases, a peak in ***I***_***d***_***(t)*** appears. The larger the ***K***_***SI***_, the higher the peak and the earlier it appears. The peak in the curve indicates the point at which the rate of recovery that takes place after ***t***_***r***_ days; plus, the rate of transfer to pool ***A*** equals the rate of new infections. The plots of ***D, A*** and ***R***_***d***_ concur with this behaviour and follow from it. ***A*** has a shape that is similar to that of ***I***_***d***_***(t)***, but at a substantially lower level.

In Fig. 5, the plots of the undetected, infected pools ***I***_***nd***_ and ***R***_***nd***_ are shown for the same values of ***K***_***SI***_ as in Fig. 4 above. Their plots are similar to those of ***I***_***d***_***(t)*** and ***R***_***d***_***(t)***, respectively. The calculation of those values is only possible if the knowledge of ***ϕ*** is accessible from public seroprevalence data.

**Figure 5.**
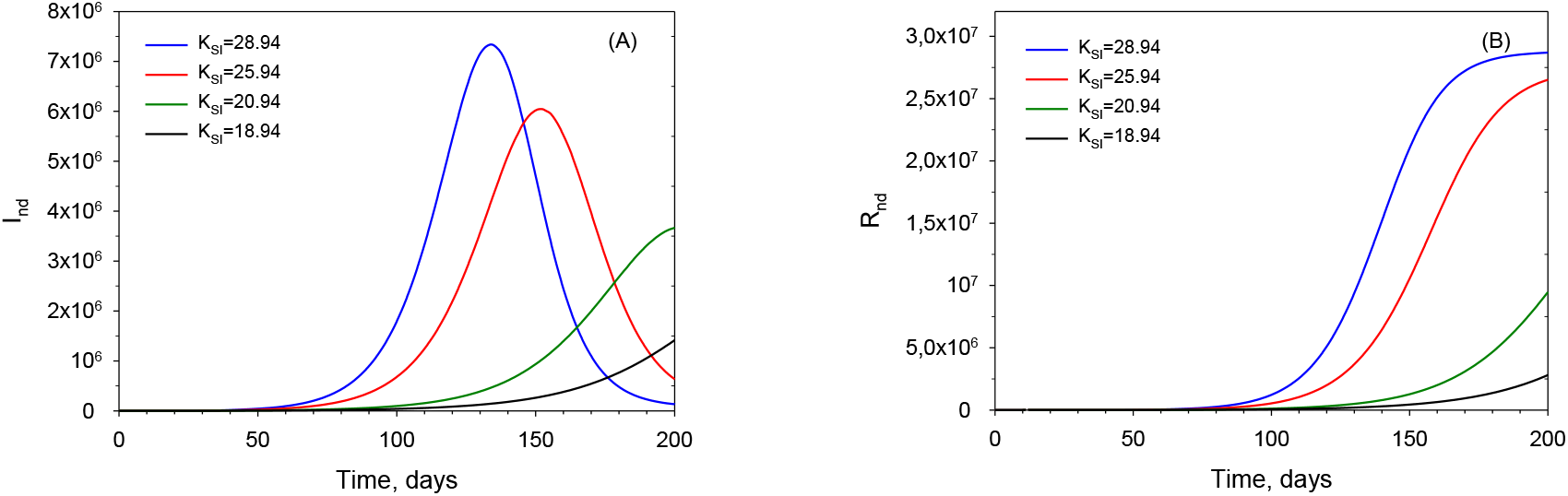
Calculated plots of undetected, infected (***A***), and undetected, recuperated (***B***) for increasing values of the basic infectivity coefficient, ***K***_***SI***_.

## 5. Quarantine

The quarantine that is usually applied in most countries may be incorporated into the present model. Every time a patient is identified as having COVID-19, an epidemiologic study is carried out and all persons suspected of having been in potentially contagious contact with him/her are sent into quarantine, isolated for a duration that is usually less than ***t***_***r***_. Those persons are indicated in the integrated scheme shown in Fig. 6, as pool ***Q***. Those who test positive for COVID-19, during or at the end of the quarantine period, go to the ***I***_***d***_ pool, while the rest rejoin the susceptible in the ***S*** pool.

**Figure 6.**
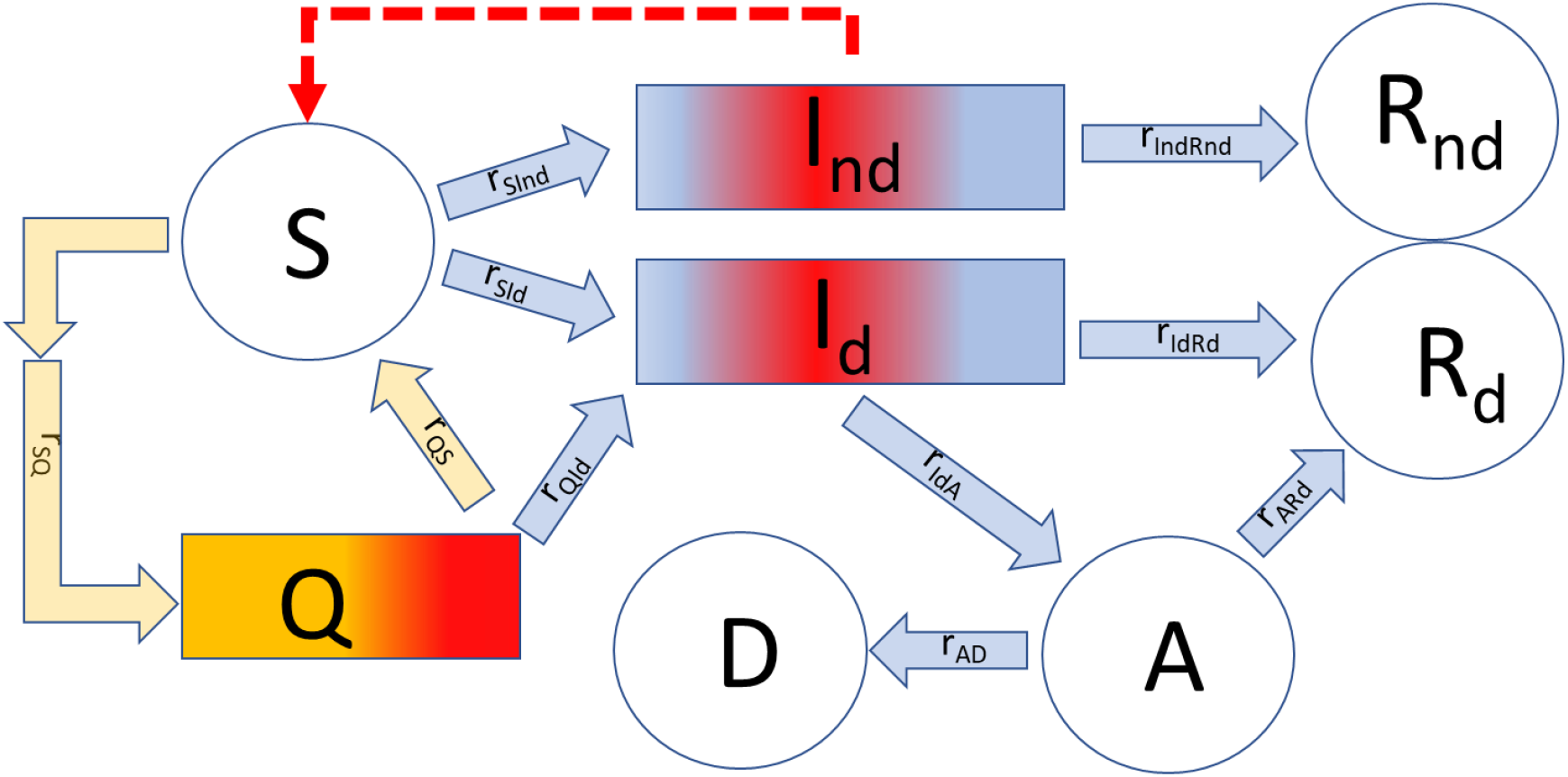
Integrated scheme of the Infection Units Model including the quarantine pool, ***Q***. The main associated parameter is the number of isolated persons per one confirmed infected, ***w***.

Strictly, those ***Q*** individuals usually enter the ***I***_***d***_ pool at a known ‘residence age’ larger than zero; this is, however, ignored in the present version of the model for the sake of simplicity. Additionally, precise data on the number of persons under quarantine are scant and not very reliable. Note that, here, ***Q*** is not the latent or pre-symptomatic stage that has been defined in other sources, but represents the actual quarantine as conducted in most countries. Quarantined individuals exit the ***S*** pool, at least temporarily, when joining the ***Q*** pool. The procedure of release from quarantine has several variants that may include the simple requirement of the asymptomatic passage of a certain amount of time and/or one or two negative COVID-19 tests; those testing positive enter pool ***I***_***d***_(t), while the rest rejoin the ***S*** pool. For the present demonstration and for the sake of simplicity, we assume that the average ‘residence time’ (***t***_***r***_) of an individual in pool Q (***t***_***rQ***_), until receiving the results of the COVID-19 test from the health authorities, is **0.3·*t***_***r***_. We represent the number of isolated individuals per one confirmed infected by the letter ***w***.

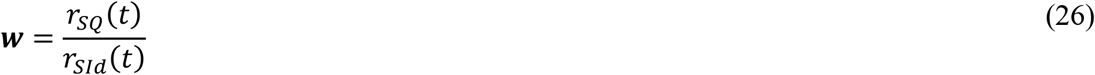

In order to reformulate the model to integrate pool **Q**, we rewrite Eqs. (12), HYPERLINK \l “bookmark10” (13), (23) and (24) as:

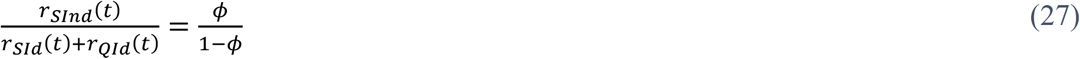

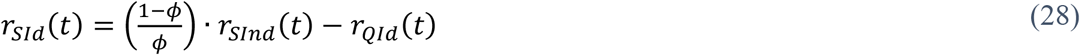

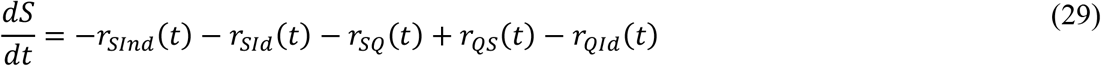

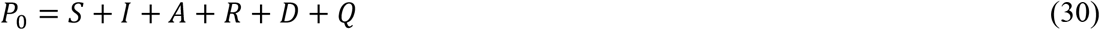

In Eq. (29), the first and the second terms on the right-hand side correspond to the outputs from pool ***S*** to the two pools of infected people, ***I***_***nd***_(***t***) and ***I***_***d***_(***t***), respectively, as seen in Eq. (23). The third term in Eq. (29), ***r***_***SQ***_(***t***), represents the flux from pool ***S*** to ***Q***, which is simply ***w*** times the flux from ***S(t)*** to ***I***_***d***_(***t***). The ratio ***w*** is defined in Eq. (31) below:

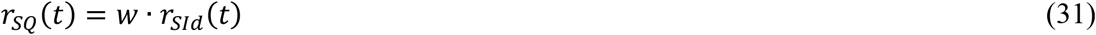

The fourth and fifth terms are related to the exit of individuals from pool ***Q***. The fourth term, ***r***_***QS***_(***t*)** is the net output from pool ***Q*** to ***S***, defined as follows:

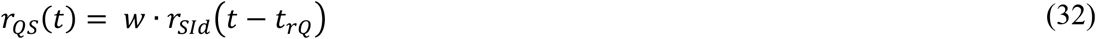

It has been assumed that this flux is proportional to the number of persons in isolation, ***Q***. The term ***r***_***QId***_(*t*) is given by:

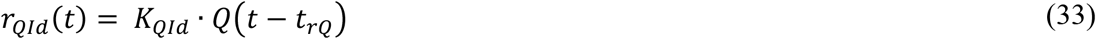

Figure 7 shows the influence of ***w*** on the ***I***_***d***_ plot. A higher value of ***w*** attests to the more stringent prophylactic measures that were taken. Thus, this model stresses the importance of quarantine in the management and control of the pandemics.

**Figure 7.**
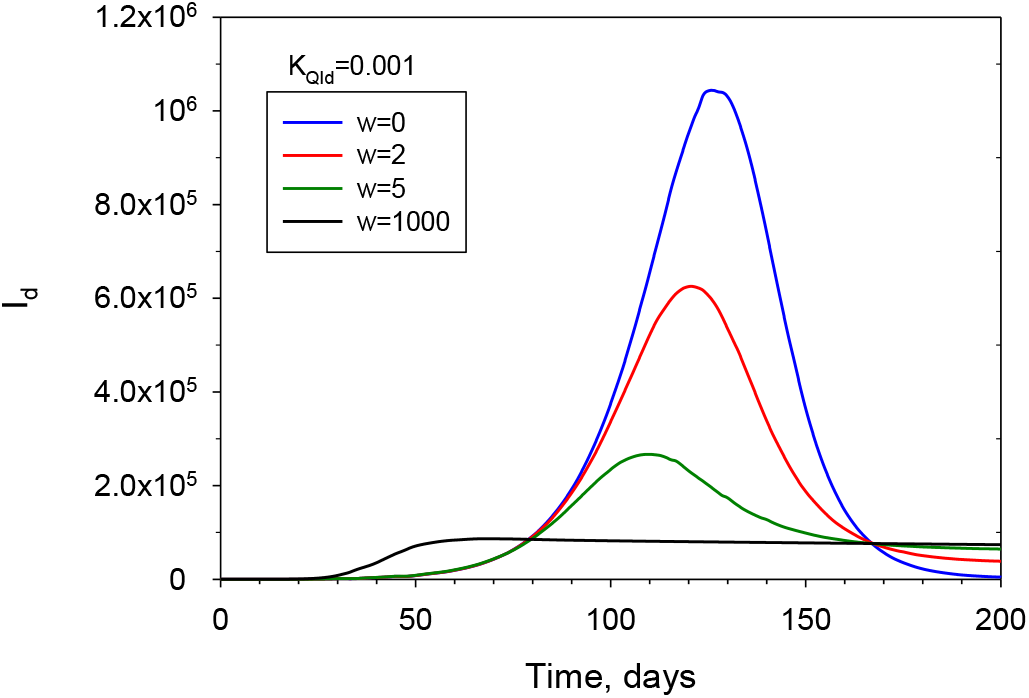
Calculated plots of diagnosed, infected (***I***_***d***_) for increasing values of the mean number of isolated persons per one confirmed infected (***w***).

## 6. Conclusions

A novel, deterministic mathematical model of the spread of the COVID-19 virus is presented here. This model describes the mechanism of the spread of pandemics, as it is seen in practice. It includes variables such as the number of individuals in pools: ***S*** (susceptible); ***I***_***d***_ (diagnosed, infected); ***I***_***nd***_ (undetected, infected); ***I***_***A***_ (assisted, hospitalized); ***I***_***dR***_ (diagnosed,recuperated); ***I***_***ndR***_ (undetected, recuperated); and of the ***Q*** (quarantined) individuals in a given population pool. The main novelty in our model consists of presenting of the ***I*** pool, <u>not</u> as a mixed compartment, but as a plug flow system, in which each individual remains for a fixed period of time. In this sense the present model is conceptually different from *SIRD*-type models and, in fact, from most of the available deterministic models. It is expected that the key ideas in this model will become useful building-blocks for the construction of more complex and generalized ‘supermodels’, that should include metapopulations, agent-based elements, statistical tools, cultural and economic aspects and their influence on the spread of pandemics, such as COVID-19.

## 7. Nomenclature

***A*** Assisted, hospitalized individuals

***a(t)*** Function representing influence of restrictions imposed by ‘social distancing’ on social contact (dimensionless)

***b*** Parameter in the ***r***_***u***_ equation which modulates the width of the shifted ***r***_***u***_ Gaussian curve (days)

***D*** Dead individuals

***f*** Encounter function (dimensionless)

***f***_***o***_ Coefficient in the encounter function ***f*** (dimensionless)

***I*** Infected individuals

***I***_***d***_ Diagnosed infected individuals

***I***_***nd***_ Undetected infected individuals

***K***_***AD***_ Rate constant in ***r***_***AD***_ (day^-1^)

***K***_***ARd***_ Rate constant in ***r***_***ARd***_ (day^-1^)

***K***_***IdA***_ Rate constant in ***r***_***IdA***_ (day^-1^)

***K***_***IdRd***_ Rate constant in ***r***_***IdRd***_ (day^-1^)

***K***_***QId***_ Rate constant in ***r***_***QId***_ (day^-1^)

***K***_***SI***_ Infectivity coefficient in ***r***_***SI***_ (infected persons ***U***^-1^)

***P***_***0***_ Target population (persons)

***Q*** Isolated individuals

***R*** Recovered individuals

***r***_***AD***_ Flux of individuals from pools ***A*** to ***D*** (persons/day^-1^)

***r***_***ARd***_ Flux of individuals from pools ***A*** to ***R***_***d***_ (persons/day^-1^)

***R***_***d***_ Recovered individuals from pool ***I***_***d***_ (diagnosed, infected persons)

***r***_***IdA***_ Flux of individuals from pools ***I***_***d***_ to ***A*** (persons/day^-1^)

***r***_***IdRd***_ Flux of individuals from pools ***I***_***d***_ to ***R***_***d***_ (persons/day^-1^)

***r***_***IndRnd***_ Flux of individuals from pools ***I***_***nd***_ to ***R***_***nd***_ (persons/day^-1^)

***R***_***nd***_ Recovered individuals from pool ***I***_***nd***_ (undetected, infected persons)

***r***_***QId***_ Flux of individuals from pools ***Q*** to ***I***_***d***_ (persons/day^-1^)

***r***_***QS***_ Flux of individuals from pools ***Q*** to ***S*** (persons/day^-1^)

***r***_***SI***_ Flux of individuals from pools ***S*** to ***I*** (persons/day^-1^)

***r***_***Sid***_ Flux of individuals from pools ***S*** to ***I***_***d***_ (persons/day^-1^)

***r***_***Sind***_ Flux of individuals from pools ***S*** to ***I***_***nd***_ (persons/day^-1^)

***r***_***SQ***_ Flux of individuals from pools ***S*** to ***Q*** (persons/day^-1^)

***r***_***u***_ Individual infectivity of an infected person (***U/***day^-1^/individual^-1^)

***r***_***umax***_ Maximum value of the ***r***_***u***_ equation (=1) (***U*** day^-1^ individual^-1^)

***S*** Susceptible individuals

***t*** Time (day)

t_c_ Starting time of restriction implementation by authorities (day)

***t***_***r***_ Serial interval - average ‘residence time’/’age’ in the ***I*** pool (day)

***t***_***rQ***_ Average ‘residence time’/’age’ in the ***Q*** pool (day)

***U*** Infection units (dimensionless)

Greek letters

***w*** Mean number of isolated persons per one confirmed infected (persons)

***β***_***f***_ The limit value towards which *a*(*t*) tends (dimensionless)

***β***_***o***_ The starting value of the function *a*(*t*) (dimensionless)

***λ*** Decay rate of social contact (day^-1^)

***τ*** Dimensionless residence time, ***t***_***r***_, defined in the ***I*** pool (-)

***τ***_***IdA***_ Typical age of the patient passing from pool ***I***_***d***_ to that pool ***A*** (-)

***τ***_***max***_ Point in **τ** where ***r***_***umax***_ is reached in the ***r***_***u***_ equation (-)

***τ***_***mix***_ Point in **τ** where ***r***_***umin***_ is reached in the ***r***_***u***_ equation (-)

***ϕ*** Fraction of infected individuals (***I***) that were undetected, **I**_**nd**_ (dimensionless)

***Ψ’***_***nd***_ Infectivity of the ***I***_***nd***_ pool for ***t*<*t***_***r***_ (***U*** day^-1^)

***Ψ***_***nd***_ Infectivity of the ***I***_***nd***_ pool for ***t*≥*t***_***r***_ (***U*** day^-1^)

## Data Availability

The relevant data are included in the MS

## Notes

### Competing Interest Statement

The authors have declared no competing interest.

